# Results Availability and Timeliness of Registered COVID-19 Clinical Trials: A Cross-Sectional Study

**DOI:** 10.1101/2021.04.07.21255071

**Authors:** Maia Salholz-Hillel, Peter Grabitz, Molly Pugh-Jones, Daniel Strech, Nicholas J. DeVito

**Affiliations:** QUEST Center for Transforming Biomedical Research, Berlin Institute of Health (BIH) at Charité – Universitätsmedizin Berlin, Berlin, Germany; The DataLab, Nuffield Department of Primary Care Health Sciences, University of Oxford, Radcliffe Observatory Quarter, Woodstock Rd., Oxford, OX2 6GG, United Kingdom

## Abstract

**Objective:** To examine how and when the results of COVID-19 clinical trials are disseminated.

**Design:** Cross-sectional bibliographic study

**Setting:** The COVID-19 clinical trial landscape

**Participants:** 285 registered interventional clinical trials for the treatment and prevention of COVID-19 completed by 30 June 2020

**Main outcome measures:** Overall reporting and reporting by dissemination route (i.e., by journal article, preprint, or results on a registry); time to reporting by dissemination route.

**Results:** Following automated and manual searches of the COVID-19 literature, we located 41 trials (14%) with results spread across 47 individual results publications published by 15 August 2020. The most common dissemination route was preprints (n = 25) followed by journal articles (n = 18), and results on a registry (n = 2). Of these, four trials were available as both a preprint and journal publication. The cumulative incidence of any reporting surpassed 20% at 119 days from completion. Sensitivity analyses using alternate dates available and definitions of results did not appreciably change the reporting percentage. Expanding minimum follow-up time to 3 months increased the overall reporting percentage to 19%.

**Conclusion:** COVID-19 trials completed during the first six months of the pandemic did not consistently yield rapid results in the literature or on clinical trial registries. Our findings suggest that the COVID-19 response may be seeing quicker results disclosure compared to non-emergency conditions. Issues with the reliability and timeliness of trial registration data may impact our estimates. Ensuring registry data is accurate should be a priority for the research community during a pandemic. Data collection is underway for Phase 2 of the DIRECCT study expanding our trial population to those completed anytime in 2020.

## INTRODUCTION

Clinical trials drive evidence generation in medicine. Clinical trial registries support transparency, accountability, and reducing bias in clinical research. The importance of these functions has been amplified during the COVID-19 pandemic. As global research efforts ramped up to address how best to treat and prevent infection with Sars-CoV-2, registrations became crucial to informing how the academic community discussed and anticipated the trajectory of clinical advancements. The rapid growth in COVID-19 research is notable: on 25 March 2020, the World Health Organisation (WHO) International Clinical Trials Registry Platform (ICTRP) listed 668 registered COVID-19 studies; on 26 March 2021 that number had grown to 8798 studies. Similarly, over 3500 reviews of COVID-19 studies in humans are registered on the PROSPERO database as of March 2021.[1]

Expectations for sharing of trial results changes during public health emergencies. The World Health Organisation (WHO) noted in a 2015 consultation that “every researcher that engages in generation of information related to a public health emergency or acute public health event with the potential to progress to an emergency has the fundamental moral obligation to share preliminary results once they are adequately quality controlled for release.”[2–4] The WHO statement did not specify an exact timeframe but rather expressed that the usual expectation to report primary results within 12 months “should be greatly shortened.” Every public health emergency is unique and brings its own realities as to how, when, and where results may become available; however, the need for rapid yet accurate dissemination remains.

Prior work on how trial results are shared in public health emergencies is sparse. Investigations of the 2009 H1N1 pandemic showed that just 29% of randomized trials examining H1N1 were reported in the literature within 18 months of trial completion, and just 12% of H1N1 vaccine trials published within a year of completion, rising to 30% within two years.[5,6] Results availability for trials during other recent major infectious disease outbreaks, for instance Ebola, Zika, MERS, or SARS-CoV-1, do not appear to have been examined to date. However, the global scale and scope of the COVID-19 pandemic, and its accompanying explosion in clinical research, distinguishes it from other modern disease outbreaks.[7] The rapid rise in the usage of preprint servers for the clinical and biomedical sciences is also a distinct feature of the current pandemic.[8–11]

We therefore set out to examine results dissemination of registered clinical trials during the COVID-19 pandemic. We aimed to explore when and where detailed trial results are being made publicly available. Understanding how and when results are shared in response to international demands for rapid, accurate data can help inform how the scientific community plans for and approaches future public health crises.

## METHODS

The DIssemination of REgistered COVID-19 Clinical Trials (DIRECCT) study was designed as a multi-phase, living examination of results dissemination throughout the COVID-19 pandemic. Phase 1 of DIRECCT is part of the CEOsys (COVID-19 Evidence Ecosystem) project funded within the Network of University Medicine (Nationales Forschungsnetzwerk der Universitätsmedizin - NUM) by the Federal Ministry of Education and Research of Germany (Bundesministerium für Bildung und Forschung - BMBF). Here we examine trials completed during the first six months of the pandemic (i.e., through 30 June 2020). We plan to expand our results in two subsequent phases covering trials completed during the pandemic’s first 12 and 18 months. Additional outcomes will be reported following final data collection. Phase 2 of the project is currently underway. The protocol for this study, with additional methods details, was preregistered on the OSF (https://osf.io/26yuj/).

### Trial Population

The World Health Organization (WHO) International Clinical Trials Registry Platform (ICTRP) maintains a list of registered COVID-19 studies across 18 global registries (Table 1). This database was downloaded on 1 July 2020 (last updated 29 June 2020) and processed to clean and standardise certain data elements across various registries (e.g., phase, study type). Known cross-registrations, taken from the COVID-19 TrialsTracker project (run by NJD), were collapsed to single trial record, preferring a ClinicalTrials.gov entry when available, unless another registration is deemed more complete by the study team, per protocol.

**Table 1:** ICTRP Data Provider Registries.

| **Registry Name** | **Abbreviation** |
| --- | --- |
| Australian New Zealand Clinical Trials Registry | ANZCTR |
| Brazilian Clinical Trials Registry | ReBec |
| Chinese Clinical Trial Registry | ChiCTR |
| ClinicalTrials.gov (USA) | N/A |
| Clinical Research Information Service (South Korea) | CRiS |
| Clinical Trials Registry - India | CTRI |
| Cuban Public Registry of Clinical Trials | RPCEC |
| EU Clinical Trials Register | EUCTR |
| German Clinical Trials Register | DRKS |
| Iranian Registry of Clinical Trials | IRCT |
| ISRCTN | ISRCTN |
| Japan Primary Registries Network* | JPRN |
| Lebanese Clinical Trials Registry | LBCTR |
| Thai Clinical Trials Registry | TCTR |
| The Netherland National Trial Register | NTR |
| Pan African Clinical Trial Registry | PACTR |
| Peruvian Clinical Trial Registry | REPEC |
| Sri Lanka Clinical Trials Registry | SLCTR |

\*Contains data from the JapicCTI, JMACCT CTR, jRCT, and UMIN CTR registries

### Inclusion/Exclusion Criteria

Inclusion and exclusion criteria for included trials were assessed twice. First using automated methods on data fields extracted directly from the ICTRP and registries between 30 June 2020 and 5 July 2020, and then via a manual assessment of each trial passing automated screening. We note two additional *post hoc* exclusion criteria that arose from coding reconciliation discussions.

Inclusion Criteria:

- Trial assessed an intervention for treatment or prevention of COVID-19 infection and subsequent acute disease.
- Trial included a completion date, or primary completion date, on or before 30 June 2020.

Exclusion Criteria:

- Registration was found, at any time, to indicate that the trial was withdrawn before enrollment and therefore never occurred.
- Registration prior to 1 January 2020.
- Trial on symptomatic treatment of COVID-19 disease side-effects only (*post hoc*)
- Trial on rehabilitation after acute disease (*post hoc*)

### Search Strategy

#### Automated Searches

On 30 June 2020, we searched PubMed using a PRESS Peer Reviewed search strategy for COVID-19 trial publications from the COVID-evidence project [12] and downloaded the XML records for all results. Next we downloaded the CORD-19 database which includes full-text and metadata for a comprehensive collection of open access coronavirus-related literature, including the PubMed Commons (PMC).[13,14] We limited both collections to only those publications as of 1 Jan 2020. We further limited our sample to publications that either (1) matched a regular expression pattern for a ICTRP-approved primary registry ID, prefix, and/or name in either the abstract, metadata, or full text (for the CORD-19/PMC sample only), or (2) was designated as a clinical trial “publication type” in PubMed. Following de-duplication, all hits were screened by two reviewers to determine whether they represented *bona fide* trial results.

#### Manual Searches

Following automated exclusion of non-interventional, pre-2020, and non-completed clinical trials, we manually reviewed all remaining clinical trials to assess their inclusion status. If a trial was included, we adapted a search strategy from Wieschowski and colleagues[15] and searched all registry entries along with PubMed, Europe PMC, Google Scholar, and Google in a stepwise fashion for results. We searched all databases using, first, the trial ID(s) and, second, keywords derived from the trial registration. Per protocol, each searcher built keyword searches using the title/abbreviation of the trial, investigator names and affiliations, and the intervention/population under study; searchers also had discretion to search additional relevant keywords from the registry as appropriate. Searches were further filtered by date ranges as necessary, and the first 30 results of a given database were checked for relevant results. All searches took place between 21 October 2020 and 18 January 2021. Once a full-results preprint and journal publication were located for a given record, no additional searches beyond a review of the registry entry were performed. All searches were performed by at least two reviewers. For each manual search, we recorded any results located, the date of publication, and whether the publication contained a “last patient, last visit” date for the primary outcome. Discrepancies were resolved by consensus between either the two reviewers or the full study team.

### Outcomes

#### Outcome Definitions

When a results publication was located we recorded it as either a journal publication, preprint publication, registry result, or other type of result. Other results types (e.g., secondary analyses, conference abstracts, grey literature) were recorded for completeness of our dataset but not included in any outcome assessments. In addition, we recorded whether the publication included the complete results of the primary outcome(s) for all participants, or if the results were interim results without complete follow-up. The primary outcome was assessed as described in the publication, and not compared to the registered primary outcome(s). Our main results are based on publication of complete, non-interim results.

#### Outcome Reporting

Per protocol, our outcome assessments are based on registry data as it stood on 30 June 2020. Subsequent updates to the completion date are not reflected in our main analysis. The only exception is if a results publication clearly included a specific “last patient, last visit” date that contradicted the registered completion date. If this occurred, we used the published completion date in all analyses.

For all trials included in our per-protocol analysis, we report overall results availability in any format by 15 August 2020 (6 weeks after our cutoff) and each specific dissemination route. We also report these results broken down by registry and by trial country. Additionally, we fit cumulative incidence curves using the Kaplan-Meier method for time to any results availability and time to journal publication. Cumulative incidence for time to preprint publication was fit using the Aalen-Johansen method with journal publication as a competing risk and ties broken by nominal offsets.[16] For trials with multiple publications, we selected the earliest publication date for the dissemination route(s) relevant to each analysis. We separately calculated the median and IQR for time to publication among only those trials with a result.

#### Sensitivity Analyses

Following data collection, we assessed the robustness of our per-protocol analyses to changes in completion date and results definitions. We assessed how the overall reporting rate changes when we: used completion dates extracted from our later manual searches rather than from 30 June 2020; used full completion dates rather than primary completion dates when available; and expanded our definition of results to include interim results. As our searches did not start until October 2020, we can also incidentally report results availability beyond our six-week minimum follow-up cutoff.

### Protocol Deviations

In addition to the *post hoc* sensitivity analyses described above, we report the following protocol deviations. We decided to delay manual searches in order to provide all completed trials at least 6 weeks to report (i.e., by 15 August 2020). This exact cutoff was not specified in our protocol. For our first round of searches, we began searches 15 weeks following our cutoff due to extensive development and piloting of our search and data extraction methods. We plan to maintain this 6-week search buffer as a minimum for results searches moving forward.

We also noted that we would continuously check inter-rater reliability for each reviewer pair. This was difficult and impractical to implement into our eventual workflow for assessing results availability. Therefore we developed an alternate system in which each reviewer-pair would attempt to reconcile any differences on study inclusion or results availability and categorisation through consensus. If consensus could not be reached, the issue was referred to the larger study team for discussion, a final consensus decision, and if necessary a specific rule addressing similar situations moving forward. We created two *post hoc* exclusion criteria based on these adjudications as detailed above. For future data extraction tasks (e.g., determining directionality of results, extracting intervention information) we may reintroduce inter-rater reliability calculations as necessary.

### Software, Code, and Data

ICTRP and registry data collection, management, and parts of the analysis were performed in Python 3.8.1 (Python Software Foundation). Additional data preparation and analysis was performed in R version 4.0.2 (R Foundation for Statistical Computing). All code and data for this project is available on GitHub.[17,18] Manual data extraction was conducted via Qualtrics,[19] and reviewer reconciliation via Numbat Systematic Review Manager.[20]

### Patient and Public Involvement

Patients and the public were not formally involved in developing this study.

## RESULTS

As of 30 June 2020, the ICTRP COVID-19 database contained information on 3,844 study registrations. Following all automated and manual exclusions, our final analysis dataset included 285 completed clinical trials. Details of all exclusions is available in Figure 1. Brief descriptive details of the 285 trials are included in Table 2.

**Table 2:** Characteristics of Included Trials.

|  | **Results Disseminated** |  |  |
| --- | --- | --- | --- |
| **Characteristic** | Overall, N = 285<sup>1</sup> | With Results, N = 41<sup>1</sup> | Without Results, N = 244<sup>1</sup> |
| **Target enrollment** | 86 (40, 200) | 60 (30, 127) | 90 (40, 200) |
| **Phase** |  |  |  |
| Not Applicable | 109 (38%) | 16 (39%) | 93 (38%) |
| Phase 1 | 13 (4.6%) | 1 (2.4%) | 12 (4.9%) |
| Phase 1/Phase 2 | 13 (4.6%) | 1 (2.4%) | 12 (4.9%) |
| Phase 2 | 46 (16%) | 5 (12%) | 41 (17%) |
| Phase 2/Phase 3 | 16 (5.6%) | 2 (4.9%) | 14 (5.7%) |
| Phase 3 | 38 (13%) | 8 (20%) | 30 (12%) |
| Phase 4 | 50 (18%) | 8 (20%) | 42 (17%) |
| **Registry** |  |  |  |
| Cross-registered | 22 (7.7%) | 2 (4.9%) | 20 (8.2%) |
| ClinicalTrials.gov | 134 (47%) | 18 (44%) | 116 (48%) |
| ChiCTR | 105 (37%) | 17 (41%) | 88 (36%) |
| IRCT | 9 (3.2%) | 1 (2.4%) | 8 (3.3%) |
| EudraCT | 3 (1.1%) | 0 (0%) | 3 (1.2%) |
| RPCEC | 3 (1.1%) | 1 (2.4%) | 2 (0.8%) |
| Registries with <3 trials | 9 (3.2%) | 2 (4.9%) | 7 (2.9%) |
| **Countries** |  |  |  |
| China | 130 (46%) | 20 (49%) | 110 (45%) |
| Iran | 22 (7.7%) | 1 (2.4%) | 21 (8.6%) |
| United States | 19 (6.7%) | 1 (2.4%) | 18 (7.4%) |
| Italy | 11 (3.9%) | 4 (9.8%) | 7 (2.9%) |
| Spain | 9 (3.2%) | 1 (2.4%) | 8 (3.3%) |
| Egypt | 7 (2.5%) | 0 (0%) | 7 (2.9%) |
| France | 7 (2.5%) | 0 (0%) | 7 (2.9%) |
| Multinational | 5 (1.8%) | 3 (7.3%) | 2 (0.8%) |
| Countries with <5 trials | 51 (18%) | 9 (22%) | 42 (17%) |
| No Country Given | 24 (8.4%) | 2 (4.9%) | 22 (9.0%) |
| <sup>1</sup>Median (IQR); n (%) |  |  |  |

**Figure 1:**
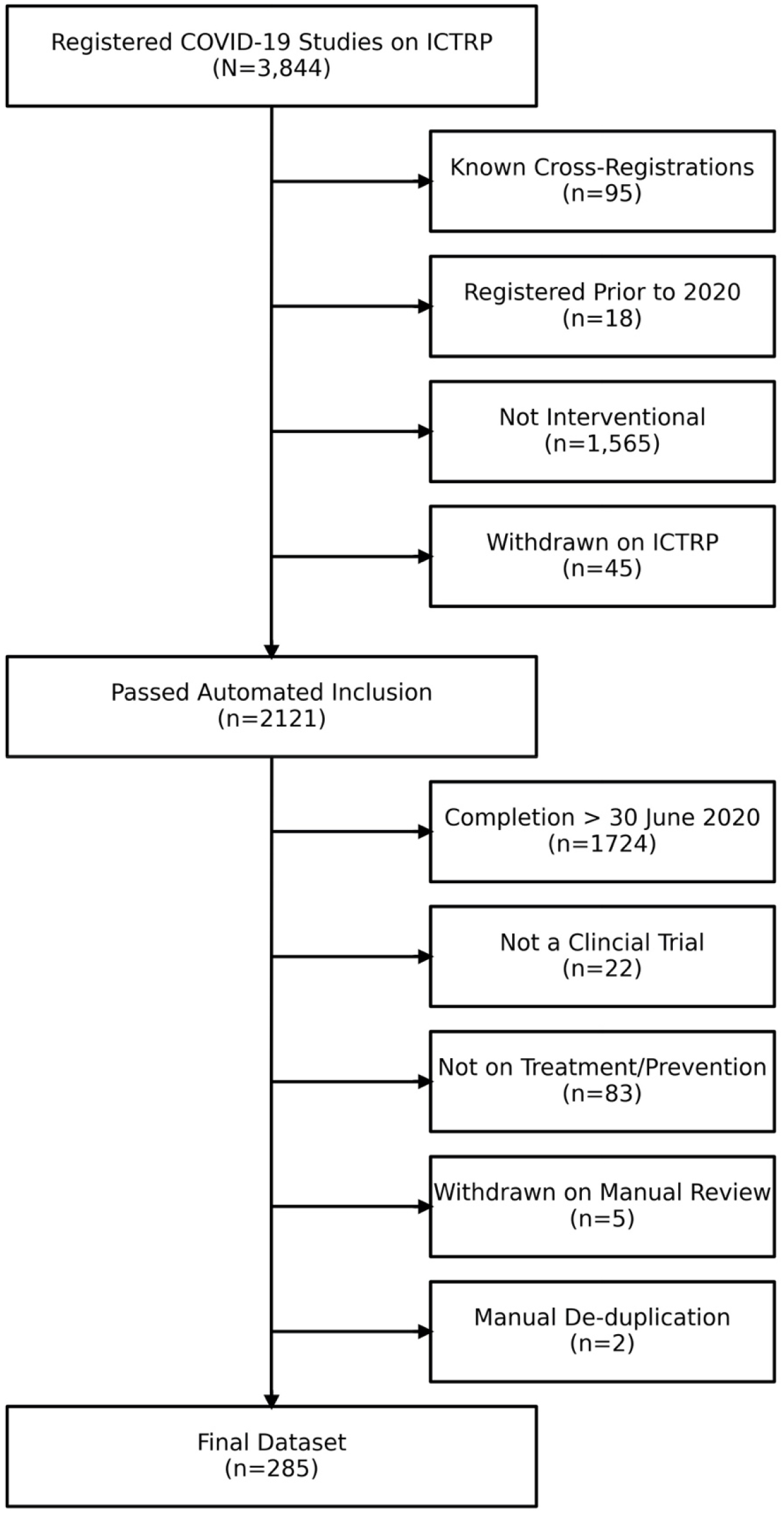
Flow Chart for Trial Inclusion.

### Per-Protocol Analysis

Among the 285 trials registered as completed by 30 June 2020, we located 41 trials (14%) with results spread across 47 individual results publications, available by 15 August 2020. Details on the screening of included results are included in Appendix A. The breakdown of results by dissemination route is detailed in Figure 2. Figure 3 shows the reporting of results by registry.

**Figure 2.**
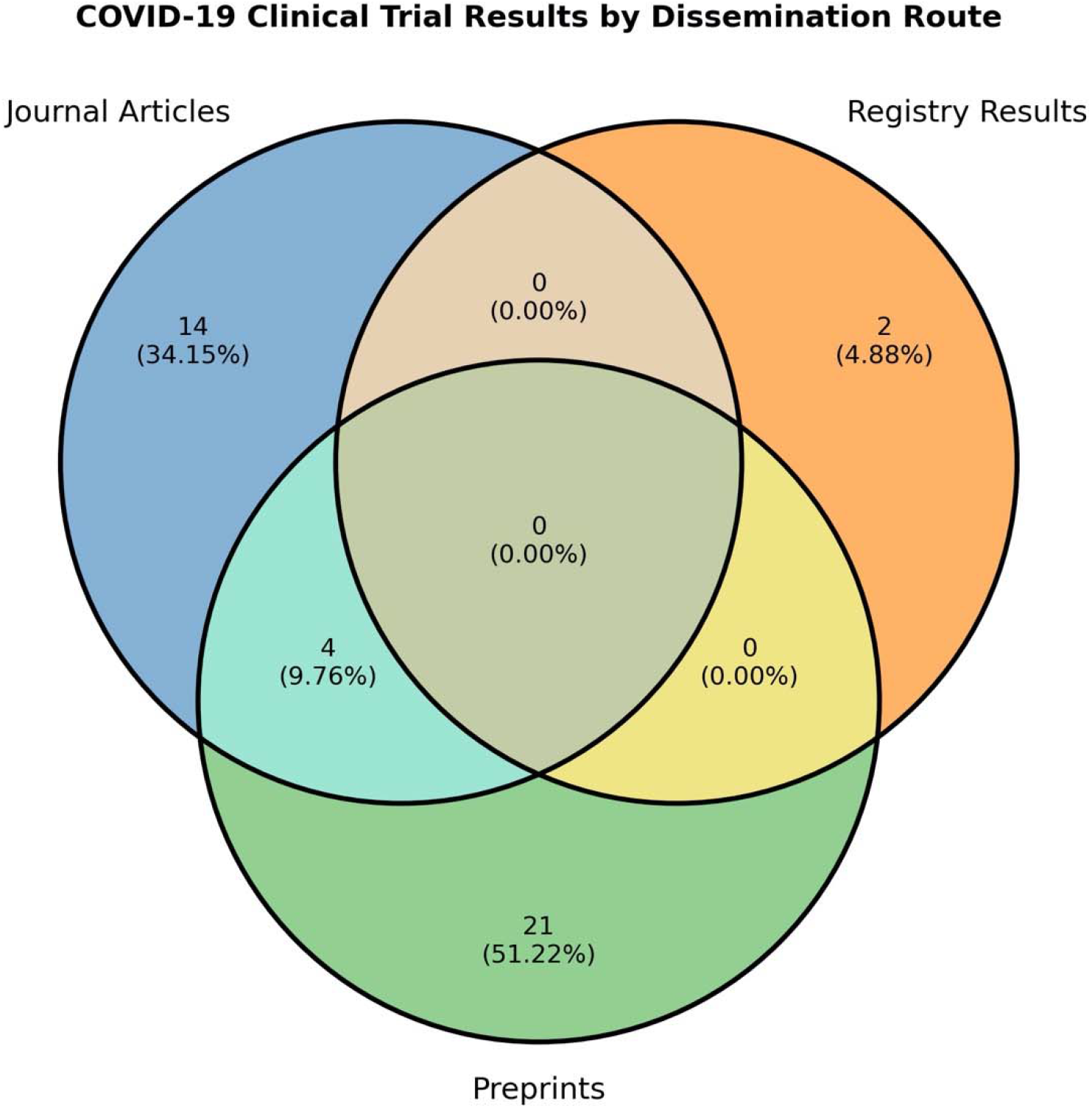

**Figure 3*.**
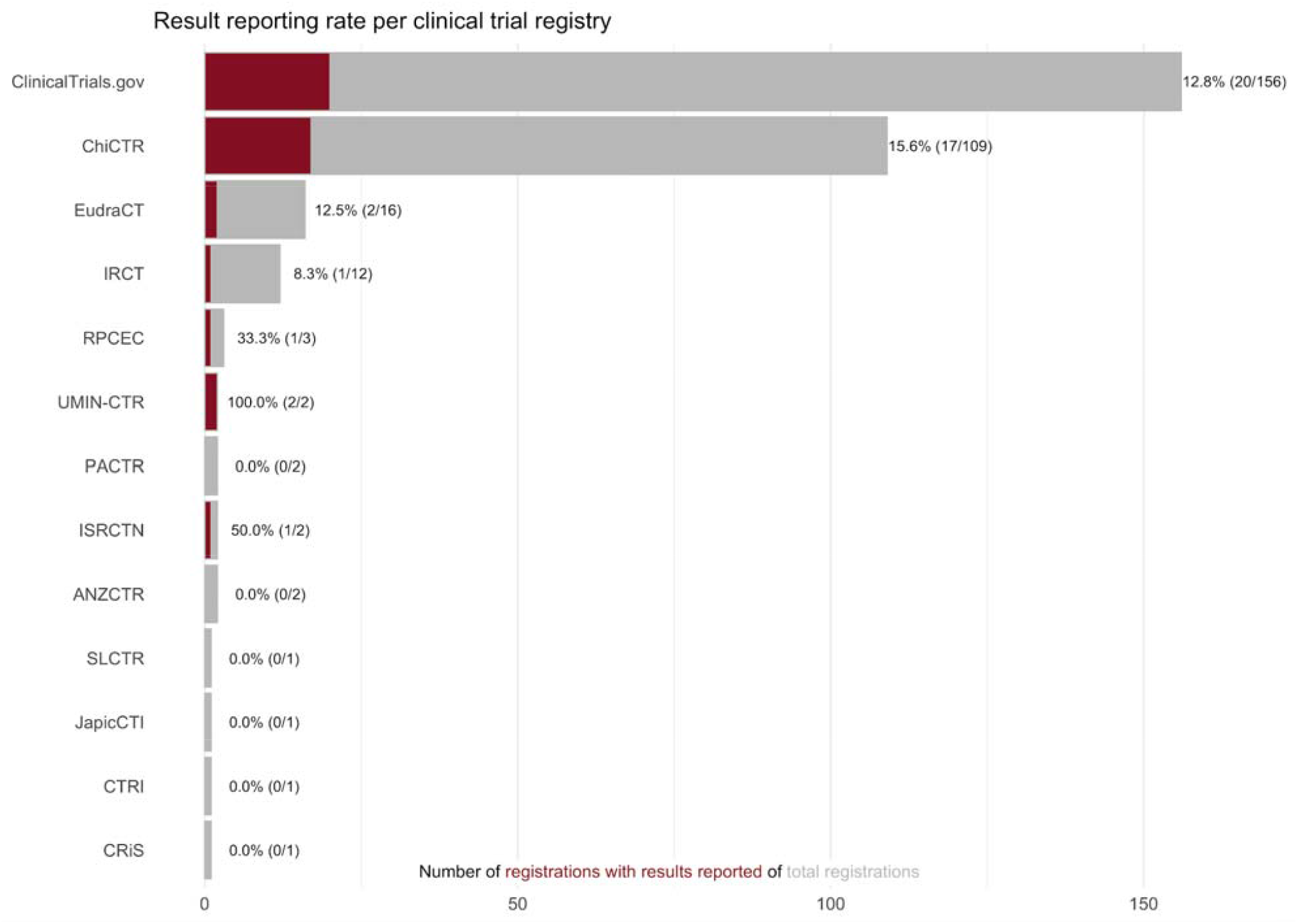
*The count of registrations is inclusive of known cross-registrations for included trials, therefore the denominators will sum to >285

Figures 4a-c shows cumulative incidence plots for time-to-publication and 95% CIs. Trials with a publication date prior to the available completion date (across all results, n = 8 of 41 trials, 19.5%) were considered reported at time 0 (except for the Aalen-Johanssen plot, which used nominal offsets to break ties at 0). If a full results publication included a different completion date (i.e., last patient, last visit), this replaced the registered completion date in the dataset (n = 4 of 41 trials, 9.8%). We show the cumulative incidence of a) first publication across any dissemination route, b) earliest journal publication, and c) earliest preprint publication. The medians for all cumulative incidence plots were undefined as no curve crossed 50%. The cumulative incidence of any results availability, accounting for censorship, surpassed 20% at 119 days from completion.

**Figure 4a-c.**
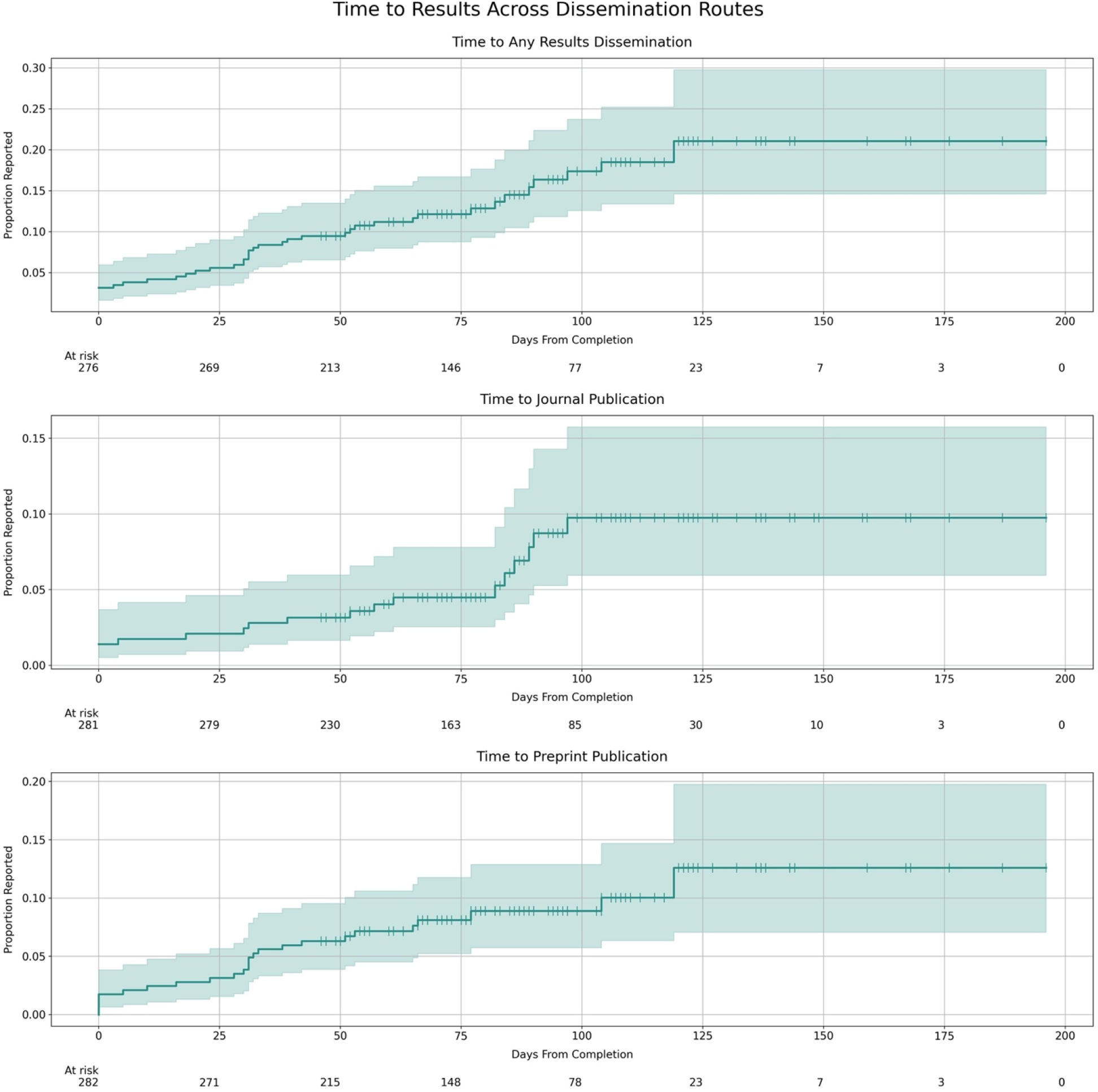

As only two trials reported results to a registry, we did not generate a plot for this route. Summary results for these two trials were published on ChiCTR 3 and 20 days following completion. For the 4 trials that had both a preprint and a journal publication, the median time from preprint publication to journal publication was 24.5 days (IQR: 14.2, 36.2). For the 21 trials that had only a preprint, preprints had been published a median of 90 days (IQR: 76, 136) by completion of follow-up.

Trials with only a preprint (n = 21) published the preprint with a median of 32 days of completion (IQR: 10, 53), whereas trials with both a preprint and a journal article (n = 4) published the preprint with a median of 31 days (IQR: 19.5, 31); trials with only a journal article (n = 14) published the article with a median of 45.5 days (IQR: 0.8, 83.5), whereas trials with both a preprint and a journal article published the article with a median of 46 days (IQR: 24.2, 67.2). Due to the small number of trials with both preprint and journal publications, we did not undertake any statistical comparisons.

### Sensitivity Analyses

Each sensitivity analysis changed only a single aspect of the main analysis, the changes were not cumulatively applied. First, we recorded a new completion date for 48 trials during manual searches, 33 of which moved the completion date post-June 2020. Using this updated trial population (n = 252) shows a reporting percentage of 16% with a minimum of 6 weeks of follow-up. Second, restricting our original sample to only those trials that reached full completion by 30 June 2020 showed 38 of 212 (18%) trials reported with a minimum of 6 weeks of follow-up. Third, adding interim results to our per-protocol findings yielded only one additional results publication for a reporting percentage of 15%.

Lastly, extending our six-week minimum follow-up window to three-months (i.e., counting any results reported by 1 October 2020), aligned with when the searches for this phase of our project actually began, adds an additional 17 results for a reporting rate of 54/285 (19%). Figure 5 includes a cumulative incidence plot for any results from expanded follow-up time.

**Figure 5.**
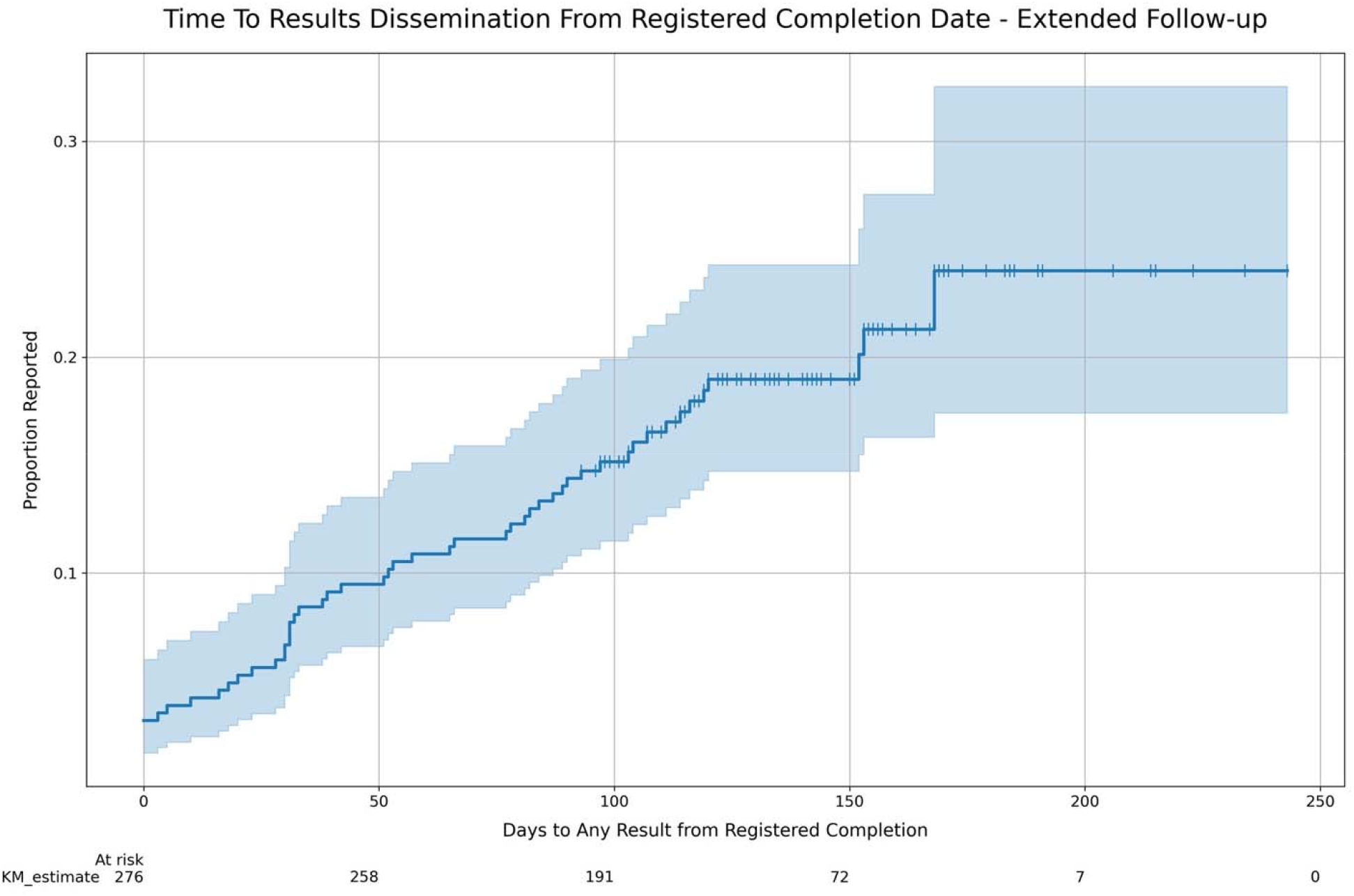

## DISCUSSION

### Summary of Findings

We identified 285 registered trials for the treatment or prevention of COVID-19 completed during the first six months of the pandemic. Of these trials, 14% had a result available in either a preprint, journal article, or posted to a clinical trials registry after a minimum follow-up time of six weeks. Sensitivity analyses using alternate dates and definitions of results for assessments did not appreciably change the reporting percentage. Extending the minimum follow-up time to three months yielded 19% of registered trials with results. Few new publications were seen beyond 120 days of follow-up. Preprints have played an important role in results dissemination with the most unique results of any route. These preprint results do not appear to be rapidly making it into the peer reviewed literature as just four preprints in our sample had a matched journal publication. There was low use (5%) of clinical trial registries for results dissemination.

### Strengths and Limitations

This work had a number of strengths. We used a broad definition of interventional clinical trials, beyond just randomized controlled trials, and included the full ICTRP database in our potential population. Searches for publications were performed using multiple strategies and databases, with dual-coded manual verification throughout. Our findings were also robust to a number of alternate assumptions for judging completion and results availability.

This work also has limitations. Missing, incomplete, inaccurate, or outdated data all likely impacted our sample to varying degrees that are difficult to quantify. Deviations from best practices for discoverability (e.g., inclusion of registration IDs in abstracts and metadata), publications in non-indexed journals or preprint servers, and articles not available in English language may have made some results difficult to locate despite our extensive search strategy. Lastly, our methods may under-represent some aspects of how results have been shared during the pandemic. For instance, large adaptive trials like the RECOVERY trial or master protocol trials like SOLIDARITY may contain numerous trials under one broad registration, produce numerous publications, and will only be represented as a single datapoint in our analysis that remains ongoing until the end of follow-up for the final arm.

### Research in Context

Registered and reported COVID-19 trials have been well-described in the literature.[7,21–25] Our results add to these examinations of the design, focus, and outcomes of the COVID-19 research response. We are aware of one similar analysis to date that attempted to quantify the dissemination of registered COVID-19 clinical trials. Janiaud and colleagues searched a single database for results of all registered COVID-19 RCTs from the first 100 days of 2020 and conducted outreach to trial contacts in October 2020. They located results for 53 of the 516 trials (10.3%) but did not match results to only registered completed trials. Of the 516 trials in their sample, 155 (30%) had not started or were discontinued, per either the registry or trial team outreach.[26]

Reporting for COVID-19 trials may be accelerated compared to other trial populations even if the overall reporting percentage remains very low. A meta-analysis from 2014 showed a pooled reporting percent of registered trials across studies of 54.2% with a minimum of 24 months of follow-up but with substantial heterogeneity. Reporting percentages in the individual studies, often examining trials in a single specialty, ranged from 23% to 76%. In five studies with time to publication data showed a 30% pooled probability of reporting at 24 months of follow-up.[27] Subsequent large examinations of reporting registered trials from academic medical centers in the US and Germany showed 35.9% and 39% of results published in the literature within two years of completion and overall reporting did not surpass 20% in either study until well after six months of follow-up.[15,28] Low reporting rates have also been seen in previous pandemics; vaccine trials during the H1N1 pandemic showed <20% probability of reporting of even within 5 months of completion.[6]

Preprints grew modestly during prior outbreaks;[8] however their rise has been substantial since the COVID-19 pandemic began, including for trials.[10,29] Preprints represented the most popular dissemination route, and often the only route, in which results were available in our sample. The low conversion of preprints to journal articles is consistent with other research on preprints during COVID-19.[30,31] The time to journal publication for preprinted trials requires additional follow-up and consideration alongside emerging evidence on the relationship between preprints and the peer-reviewed COVID-19 academic literature.[11]

### Implications for Policy and Practice

The deluge of trial registrations also reflects the fragmentation of the global COVID-19 response. As others have noted, this rush to register research without coordination or consideration of the work of others may compromise progress towards answering crucial questions.[32] A disparate research environment may lead to considerable research waste as unnecessary duplication, disjointed outcomes, and competition for participants slows evidence generation.[33–36] It is essential that we understand the COVID-19 research response and how and why it evolved as it did.

A complete assessment of COVID-19 trial results dissemination would require accurate registry data on both trial completion and the overall status of the trial. The availability of completion dates in trials considered ranged from never (e.g., REBEC), to mixed (e.g., ANZCTR), to always (e.g., ClinicalTrials.gov). Other registries, like the EUCTR, make completion dates available only retrospectively. Despite being a CONSORT element, explicit dates for trial completion are rarely included in clinical trial publications, which would aid checks for accuracy and provide context for results.[37] Issues with missing, incomplete, and outdated data on registries has been detailed in prior investigations.[38–40] Registries were also rarely used as a dissemination route for clinical trial results. The US National Institutes of Health have since called for the rapid publication of COVID-19 study results on ClinicalTrials.gov, the largest global registry, and instituted expedited reviews of results submissions moving forward.[41] Our future work will examine whether this has led to more results availability on registries.

It is difficult to conclude from these preliminary results whether our findings show true potential for publication bias from unpublished results, poor management of trial registrations, or some combination of both. The stakes of a global pandemic only amplify the importance of minimizing reporting issues that may impact evidence synthesis, guideline development, and ultimately clinical practice. Failure to update registry entries compromises an important tool for transparency and accountability in the COVID-19 research response. Searching for or anticipating results that may simply never come and/or should not be expected hampers crucial efforts to collect and examine the evidence around COVID-19.

### Future Research

Data collection for Phase 2 of the DIRECCT project is currently underway, extending our results to trials completed at any time in 2020. The continued examination of publication rates over time will help place the COVID-19 results reporting data in further context and help inform expectations for reasonable dissemination timelines in public health emergencies. In our final Phase, we plan to expand our data collection to include outreach to authors and study teams, as well as to examine factors associated with the timely reporting of COVID-19 clinical trials. Future studies may also wish to examine additional forms of dissemination, such as press releases or other grey literature findings. We hope that our openly available dataset of results matched to registrations will be useful to other groups investigating the COVID research response. Our data forms one part of a robust landscape of curation, examination, and synthesis of COVID-19 research and literature, through platforms like COVID-NMA, Cochrane, COVID-evidence, and Epistimonikos.[12,42–44] These efforts allow for rapid evidence synthesis and the ability to efficiently examine pandemic research output in order to inform strategies for improved management and coordination of the global scientific community both during global health emergencies and otherwise.

## CONCLUSION

Many of the trials registered, and apparently completed, during the first six months of the pandemic failed to yield rapid results in the literature or on clinical trial registries. Nevertheless, our findings suggest that the COVID-19 response may be seeing quicker results disclosure compared to non-emergency conditions. Issues with the accuracy and currentness of trial registration data are likely to obfuscate a more precise estimate of non-publication of clinical trials. Maintaining accuracy in clinical trial registrations should be a priority, especially during global public health emergencies, when collaboration and research prioritisation is key to the efficient advancement of knowledge. Among registered trials that did report results, preprints were the most common dissemination route, however very few had converted to a full journal publication within the timeframe of this study. Results were rarely made available on clinical trial registries. Better, more specific guidance, aligned to the real-world learnings about how, when, and where results from clinical trials should be expected during pandemic situations, may be warranted.

## Supporting information

STROBE Checklist

## Data Availability

All code for this project is available at https://github.com/ebmdatalab/covid19_results_reporting and https://github.com/maia-sh/direcct. The full dataset is available at https://www.doi.org/10.5281/zenodo.4669937.

https://github.com/maia-sh/direcct

https://www.doi.org/10.5281/zenodo.4669937

## ACKNOWLEDGEMENTS

We would like to thank Lars Hemkens (U of Basel) and Ben Goldacre (U of Oxford) for their support and advice in developing this project; Benjamin Gregory Carlisle for extensively supporting our use of the Numbat Systematic Review Manager software and making adaptations for this project; and Kerstin Rubarth from the Charité Institut für Biometrie und Klinische Epidemiologie for her consultation on the protocol and statistical analysis plan. We also thank our colleagues who provided useful comments on our study protocol and/or manuscript: James Smith (U of Oxford), Florian Naudet (CHU Rennes), and members of the DataLab at the University of Oxford and QUEST Center for Transforming Biomedical Research at the Berlin Institute of Health at Charité Universitätsmedizin zu Berlin.

## Transparency Statement

The lead author (NJD) affirms that the manuscript is an honest, accurate, and transparent account of the study being reported; that no important aspects of the study have been omitted; and that any discrepancies from the study as planned have been explained. All authors had full access to all of the data (including statistical reports and tables) in the study and can take responsibility for the integrity of the data and the accuracy of the data analysis.

## Ethical Approval

This study was exempted from ethics approval at the University of Oxford as it solely used non-disclosive, publicly available data.

## Contributorship Statement

*Study concept and design:* All authors

*Acquisition, analysis, or interpretation of data:* MSH, NJD, PG, MPJ

*Drafting of Manuscript:* NJD

*Critical revision of the manuscript for important intellectual content:* All authors

*Data curation:* MSH, NJD

*Software:* MSH, NJD

*Statistical Analysis:* NJD, MSH

*Administrative, technical, or material support:* PG, DS

*Study supervision:* DS, NJD

*Funding acquisition*: DS

*Guarantor:* NJD

## Appendix A. Screening of results publications

Through our automated and manual publication search strategies, we found a total of 124 publications of complete results as either registry results, preprints, or journal articles. 47 results were associated with trials included in our per protocol analysis. Of the 77 excluded results, 5 located during automated searches but could not be associated with a trial registration number, 8 were associated with a trial with no registered completion date, 23 were associated with a trial with a completion date following the 30 June 2020 cutoff, 1 has no publication date, 39 have a publication date following the 15 August 2020 cutoff, and 1 indicated that the trial was retrospective and was therefore post hoc excluded from our per protocol population.

**Supplementary Figure 1.**
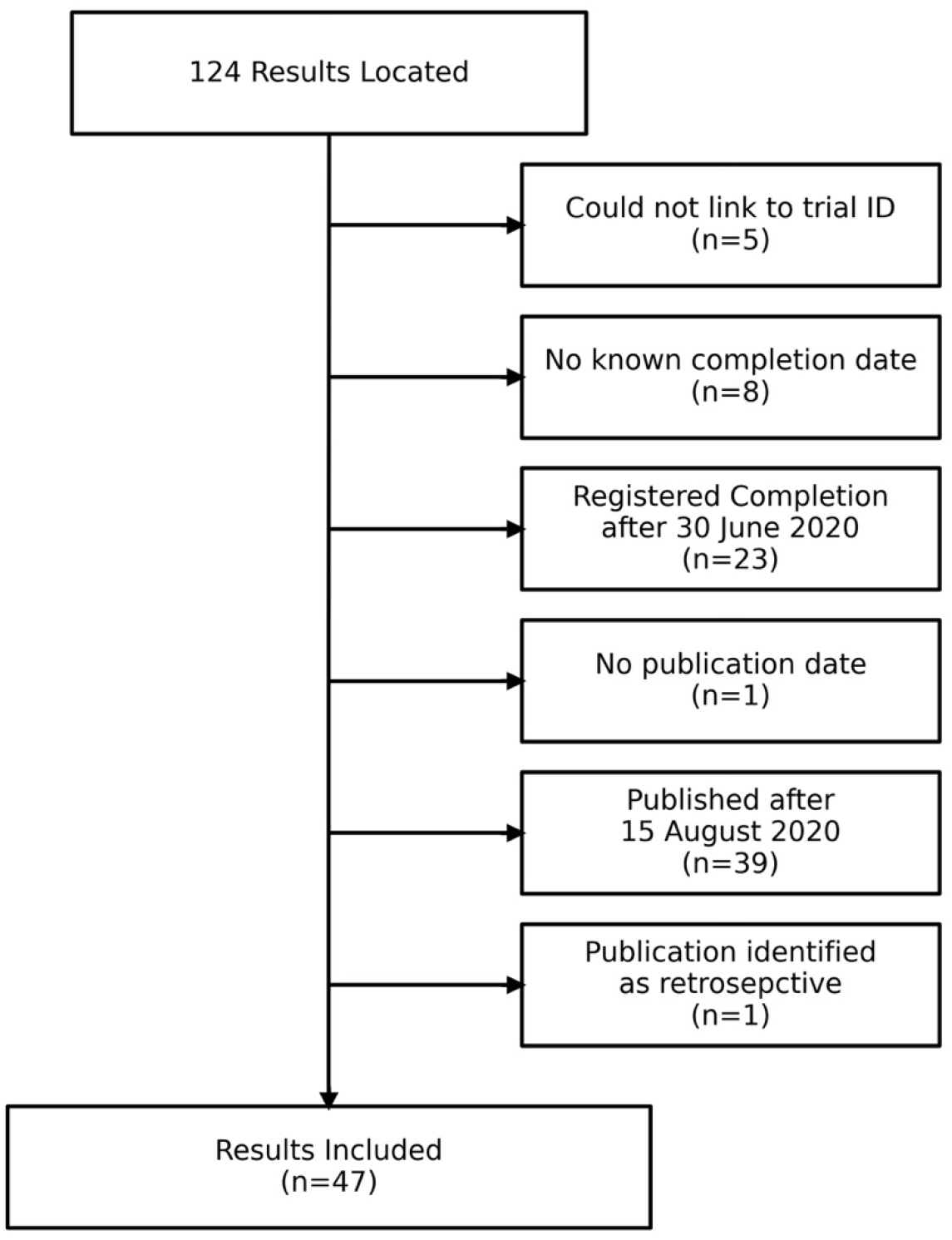
Results Flowchart.

